# Assessing the Impact of Race on Glomerular Filtration Rate (GFR) Prediction

**DOI:** 10.1101/2021.10.26.21265423

**Authors:** Linying Zhang, Lauren R. Richter, George Hripcsak

## Abstract

**Background:** The appropriate use and the implications of using variables that attempt to encode a patient’s race in clinical predictive algorithms remain unclear. The clinical algorithm for estimating glomerular filtration rate (GFR) adjusts for race, but the observed difference between Black and non-Black participants lacks biologically substantiated evidence. We investigated the impact of using a race variable on GFR prediction by race-stratified error analysis.

**Methods:** We implemented three predictive algorithms with varied amount of input information from an electronic health record database to estimate GFR. We compared the prediction error of the estimated GFR with and without the variable race between Black patients and White patients.

**Results:** The prediction error for patients coded as Black was higher than that for patients coded as White across all three algorithms. Removing race from the prediction algorithm did not lower the prediction error for patients coded as Black, neither did it decrease the difference in error between the two groups. The algorithm that included the most information with thousands of variables but excluding race produced the most accurate estimate for both groups and minimized the difference in performance between the two groups.

**Conclusion:** The prediction error for patients coded as Black was higher compared to those coded as White, regardless of inclusion of race as a variable. Using a large amount of information represented in electronic health record variables achieved a more accurate prediction of GFR and the least difference in prediction error across racial groups.

## Introduction

The appropriate use and the implications of using variables that attempt to encode a patient’s race in medical predictive algorithms remain unclear^1^. One example of an algorithm that includes a race variable is the equation for estimating glomerular filtration rate (GFR), an indicator of kidney function used to classify the severity of chronic kidney disease (CKD). The most commonly used clinical algorithm for estimating GFR, the Chronic Kidney Disease Epidemiology Collaboration (CKD-EPI) equation^2^, uses a modifier for Black race to adjust for differences in serum creatinine levels among Black research study participants as compared to their non-Black counterparts. However, the observed difference between Black and non-Black participants lacks biologically substantiated evidence^3,4^. Discussions around the ethics of including a race modifier in estimating GFR have been around for the past decade^2,5^. Recently, Diao *et al^6^* showed that removing race as a variable from the estimated GFR equation could have a significant impact on recommended care for Black patients (*e*.*g*., increasing CKD diagnoses among Black adults could improve access to specialist care and kidney transplantation). However, Diao *et al^6^* did not study whether removing the race modifier leads to more accurate GFR predictions for Black patients. The goal of our study is to investigate the impact of using a race variable on GFR prediction by race-stratified error analysis. We consider the use case of drug dosing, in which the difference between the true GFR and the estimated GFR will be relevant.

## Methods

De-identified electronic health records were collected from Columbia University Medical Center. A total of 1,560 patients (2,471 visits) were included in the cohort, of which 1,154 patients their race variable coded as White (1,835 visits) and 406 patients coded as Black (636 visits). To compare the estimated GFR (eGFR) to true GFR, we first calculated the true GFR using each patient’s 24-hour urine creatine (U_Cr_), serum creatinine (S_Cr_) and body surface area (BSA) as follows:

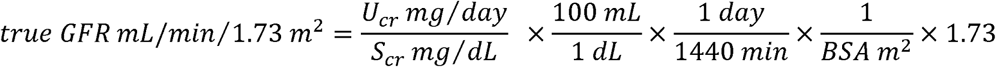

The eGFR was computed using three methods with and without the variable race.

1. The CDK-EPI equation, which is

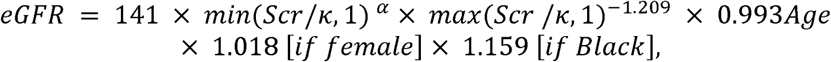

where α is − 0.329 for females and − 0.411 for males, and *κ* s 0.7 for females and 0.9 for males. To compute eGFR without the race variable using the CKD-EPI equation, the coefficient for Black race, 1.159, was removed.
2. A linear regression developed using our data with the same input variables as the CKD-EPI equation. The model is

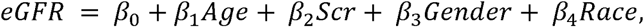

where *β*s are the coefficients, the variable gender is 1 for females and 0 for males, and the variable race is 1 for black patients and 0 for white patients. To compute eGFR without the race variable using this method, a linear regression model was trained without the variable for race.
3. an *L1*-regularized regression with all variables in the database. The model is

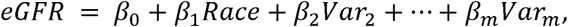

and the cost function is

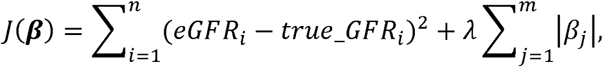

where *β*s are the coefficients and *λ* is the regularization parameter (*λ* > 0). To compute eGFR using this method without the race variable, a similar *L1*-regularized regression was trained on all variables except the variable for race.

For model 2 and model 3, in addition to training the model with and without a race variable on the entire cohort, we also trained the model on each race subgroup separately. Prediction error is defined as the root mean squared error (RMSE) between the eGFR and the true GFR on the test set. Prediction error was calculated for the entire cohort as well as on each race subgroup separately. The standard deviation is derived from 30 train-test splits. The hyperparameter is tuned by 5-fold cross-validation on each train set.

## Results and Discussion

**Table 1.** The GFR prediction error from three methods with and without a race variable. The all-covariate regression model achieved the lowest prediction error and also the least difference in error between patients coded as Black or White.

| Model |  | CKD-EPI equation | Linear regression | Regularized regression with all covariates |
| --- | --- | --- | --- | --- |
| Input | | age, sex, $S_{cr}$, +/- race | | All covariates +/- race |
|  |  | Mean RMSE (sdRMSE) |  |  |
| All patients | With race | 42.57(1.71) | 49.91(1.88) | 39.47(2.17) |
|  | Without race | 43.61(1.76) | 48.73(2.38) | 39.45(2.01) |
| Patients coded as Black | With race | 46.78(3.52) | 55.59(3.83) | 41.84(2.73) |
|  | Without race | 48.37(3.65) | 53.84(3.83) | 42.14(3.13) |
|  | Stratified by race | NA | 53.09(2.58) | 42.00(3.38) |
| Patients coded as White | With race | 40.95(2.07) | 47.70(2.45) | 38.57(2.56) |
|  | Without race | 41.77(2.17) | 46.75(3.02) | 38.40(2.60) |
|  | Stratified by race | NA | 46.53(3.56) | 38.41(2.56) |

The prediction error for patients coded as Black was higher than that for patients coded as White across all three methods. The higher prediction error for patients coded as Black could lead to inappropriate clinical decisions, and thus future research should look into how to improve prediction for Black patients. Additionally, the higher prediction error for patients coded as Black was insensitive to the use of race in the prediction algorithm. Removing race from the prediction algorithm did not lower the prediction error for patients coded as Black, neither did it decrease the difference in error between the two groups.

The model using large-scale covariates had the lowest error compared to the other two models, both with race and without race, indicating it is the best among the three methods in estimating GFR. In addition, it had the smallest gap of prediction error between the two groups, a potentially fairer algorithm if equal prediction error for all racial groups is desired. Furthermore, the regularized regression had the smallest difference of prediction error with *vs*. without race, indicating that the other covariates included in the model can potentially replace race without affecting the prediction. It is worth investigating what variables are included and what their roles might be in affecting kidney function.

## Conclusion

We did not observe a significant difference in GFR prediction error after including or excluding race in all three prediction algorithms tested. The prediction error for patients coded as Black was higher compared to those coded as White, regardless of inclusion of race as a variable. More variables other than those used in the current clinical algorithm help achieve more accurate prediction of GFR and the least difference in prediction error across racial groups.

## Data Availability

The data that support the findings of this study are not openly available due to patient privacy.

